# Individual-Level Counterfactual Analysis of SGLT2 Inhibitors Versus DPP4 Inhibitors in Diabetic Kidney Disease Using Causal Machine Learning

**DOI:** 10.64898/2026.08.30.26361750

**Authors:** Yuichiro Yano, Hajime Nagasu, Hiroshi Kanegae, Mizuki Ohashi, Yoshitaka Isaka, Hirokazu Okada, Masaomi Nangaku, Naoki Kashihara

## Abstract

**Background:** Traditional real-world studies comparing SGLT2 and DPP4 inhibitors on renal outcomes rely on propensity score matching, which causes high-dimensional data loss. We used causal machine learning (Causal ML) to unmask heterogeneous treatment effects in diabetic kidney disease (DKD).

**Methods:** Using data from 4,588 patients within the Japanese J-CKD-DB-Ex registry, we implemented a doubly robust (DR) learning framework (Linear DR-learner with XGBoost) to compare SGLT2 and DPP4 inhibitors. Outcomes included the chronic eGFR slope and a composite renal endpoint ( 50% eGFR decline or end-stage kidney disease). Heterogeneity was explored via causal SHAP and decision trees.

**Results:** At the population level, SGLT2 inhibitors modestly slowed chronic eGFR decline (average treatment effect [ATE] = 0.14 [95% CI: −0.86, 1.15] mL/min/1.73m²/year) and reduced composite endpoint risk by 9% (ATE: −0.09 [−0.11, −0.08]) versus DPP4 inhibitors. However, individual-level counterfactual analysis suggested that for the chronic eGFR slope, non-glinide users with stable pre-treatment trajectories who were also taking ACE inhibitors had a greater benefit from SGLT2 inhibitors (ATE: 2.95 [-0.68, 6.58]). Conversely, glinide users with steep pre-treatment decline had a greater benefit from DPP4 inhibitors (ATE: −8.98 [−16.11, −1.85]). For composite renal events, SGLT2 inhibitors had a 28% absolute risk reduction within the algorithmically identified high-risk subgroup (eGFR 28.1 mL/min/1.73 m² and positive proteinuria; ATE: −0.28 [−0.33, −0.23]). Even non-proteinuric decliners demonstrated a 8% risk reduction with SGLT2 inhibitors (ATE: −0.08 [−0.10, −0.06]).

**Conclusion:** Causal ML advances precision medicine in DKD, shifting from uniform prescribing to individualized, data-driven therapy targeting distinct intrarenal pathways.

## INTRODUCTION

In the management of type 2 diabetes mellitus (T2DM), preserving kidney function is a critical therapeutic goal. Recently, an accumulating body of real-world evidence (RWE) has demonstrated that sodium-glucose cotransporter 2 (SGLT2) inhibitors confer superior renoprotective effects, specifically in slowing the decline of the estimated glomerular filtration rate (eGFR) slope compared to dipeptidyl peptidase-4 (DPP4) inhibitors **[1–5]**. However, the vast majority of current RWE is based on propensity score matching (PSM) **[1–5]**. Recent methodological advancements have revealed structural and mathematical limitations of PSM in high-dimensional data spaces. The so-called PSM paradox illustrates that, under certain conditions, stringent matching can inadvertently increase imbalance, model dependence, and statistical bias by randomly discarding samples that are already naturally well-balanced **[6]**.

Causal Machine Learning (Causal ML) inherently overcomes these structural flaws of PMS by integrating the rigorous mathematical frameworks of causal inference with the high-dimensional data processing capabilities of modern ML **[7]**, including Doubly Robust (DR) learning and Neyman orthogonality. Unlike traditional PSM, which primarily focuses on average treatment effects (ATE) by comparing group means, Causal ML enables the estimation of individualized treatment effects (ITE) and conditional average treatment effects (CATE). This granular capability is essential for elucidating the comparative effectiveness of SGLT2 inhibitors versus DPP4 inhibitors in specific subgroups, such as non-proteinuric diabetic kidney disease (DKD) patients or early decliners, whose disease trajectories may differ from those with classic proteinuric nephropathy.

In this study, we applied a DR learning framework, specifically the Linear DR-learner, to evaluate the heterogeneous treatment effects (HTEs) of SGLT2 inhibitors versus DPP4 inhibitors on the eGFR slope in Japanese CKD patients with T2DM. By leveraging this advanced Causal ML approach, we aim not only to address the inherent weaknesses of PSM but also to identify the specific patient subgroups that derive the maximum renoprotective benefit from SGLT2 inhibitors, thereby informing highly personalized and effective clinical decision-making.

## METHODS

### Data Sources and Study Population

The Japan Chronic Kidney Disease Database (J-CKD-DB) is a large-scale, real-world observational database jointly developed by 21 university hospitals in Japan (UMIN trial number: UMIN000026272). Launched in December 2014, it is built on electronic health records (EHRs) encoded in the Standardized Structured Medical Information eXchange 2 (SS-MIX2) storage format. The SS-MIX2 system integrates unified coding systems for laboratory tests (JLAC10) and prescriptions **[8]**. Both inpatient and outpatient data are captured automatically via the Multipurpose Clinical Data Registry System **[9]**, which was custom-designed to extract information through SS-MIX2. An initial data extraction covering the period from January 1 to December 31, 2014, successfully minimized manual data entry, reduced physician workload, and prevented transcription errors. To ensure data integrity and interoperability, the database strictly adheres to core standards embedded in SS-MIX2, such as the HL7 V2.5 format, JLAC10 codes for laboratory results, and ICD-10 codes for diagnoses **[10]**.

Patients (≥18 years of age) were eligible for inclusion in the J-CKD-DB if they met at least one of the following criteria: (i) dipstick-positive proteinuria (≥1+), or (ii) an eGFR below 60 mL/min/1.73 m² **[11]**. This study was approved by the Institutional Review Board of Juntendo University (approval number: C25-0105) and conducted in accordance with the Declaration of Helsinki. Under Japan’s Ethical Guidelines for Medical and Health Research Involving Human Subjects, the requirement for individual informed consent was waived because all patient records were fully anonymized. Instead, an opt-out consent process was employed: patients were notified via public postings on each participating hospital’s website and provided the opportunity to withdraw their data. This procedure was explicitly authorized by the ethics committee.

### Study Design and Cohort Definition

We conducted a retrospective observational cohort study using a new-user, active-comparator design. Data were derived from the J-CKD-DB-Ex, a prospective longitudinal follow-up cohort conducted at eight university hospitals between 2014 and 2022. From this dataset, we selected records of patients with T2DM who had at least 1 year of continuous enrollment history in the database prior to initiating either an SGLT2 inhibitor or a DPP4 inhibitor. The index date was defined as the date of the first prescription for any SGLT2 inhibitor (canagliflozin, dapagliflozin, empagliflozin, ipragliflozin, luseogliflozin, or tofogliflozin) or DPP4 inhibitor (Trelagliptin, Vildagliptin, Saxagliptin, Sitagliptin, Sitagliptin, Anagliptin, Teneligliptin, Linagliptin, Alogliptin, Omarigliptin, Vildagliptin / Metformin, Alogliptin / Metformin, Anagliptin / Metformin, Alogliptin / Pioglitazone), as either initial or add-on therapy, provided no prior prescriptions for that respective medication class had been issued during the preceding year.

Consistent with established methodologies for evaluating eGFR trajectories, such as the CVD-REAL 3 study **[12]**, we included patients who had at least two eGFR measurements before the index date, with the most recent measurement occurring within 180 days prior to the index date. To ensure a reliable estimation of the pre-treatment eGFR slope, we further required an interval of at least 180 days between the first and last eGFR measurements before the index date. Patients were followed from the index date until the end of the index treatment (on-treatment analysis), migration or departure from the database, death, or the end of the data collection period, whichever occurred first.

### Kidney Function and Other Measurements and Outcomes

Serum creatinine and spot urine specimens were collected for each participant. Serum creatinine was assayed using an enzymatic method. The eGFR was calculated using the Chronic Kidney Disease Epidemiology Collaboration (CKD-EPI) equation, modified by a Japanese coefficient **[13]**. A rapid decline in kidney function prior to treatment initiation was defined as an eGFR loss of ≥3.0 mL/min/1.73 m² per year **[14]**. Urinalysis by the dipstick method was performed on spot urine specimens. Urine dipstick results were interpreted by the medical staff at each hospital and recorded as (−), (±), (1+), (2+), or (3+). According to the policy of the Japanese Committee for Clinical Laboratory Standards (https://jccls.org/), all urine dipstick tests are manufactured such that a result of 1+ corresponds to a urinary protein level of 30 mg/dL. In this study, proteinuria was defined as a dipstick result of 1+ or greater.

The primary outcome was the rate of change in eGFR (eGFR slope) following the initiation of either an SGLT2 inhibitor or a DPP4 inhibitor. Initiation of SGLT2 inhibitors frequently induces an acute, reversible decline in eGFR **[15]**. To strictly account for this acute hemodynamic phenomenon and accurately capture the true long-term trajectory of kidney function, we focused on the chronic eGFR slope. The chronic eGFR slope was calculated using eGFR measurements obtained from day 30 post-index date onwards, deliberately excluding the first 30 days of treatment. This methodology aligns with the rigorous analytical frameworks established in landmark randomized controlled trials (RCTs), including the EMPA-REG OUTCOME, CREDENCE, and DAPA-CKD trials, which routinely differentiated the initial hemodynamic dip from the chronic slope to accurately evaluate long-term renoprotective efficacy **[16–18]**. The secondary outcome was a composite endpoint consisting of a sustained reduction in eGFR of ≥50% (confirmed by a subsequent measurement) or the onset of end-stage kidney disease (ESKD), defined as an eGFR of <15 mL/min/1.73 m² (confirmed by a subsequent measurement).

### Statistical Analysis

We conducted data analyses comparing the SGLT2 inhibitor group and the DPP4 inhibitor group. Patient characteristics were summarized using frequencies and percentages for categorical variables, and means with standard deviations for continuous covariates. The balance of baseline covariates between the two groups was assessed using the standardized mean difference (SMD), with an SMD < 0.1 indicating negligible differences **[19]**. Specifically, the variables used to calculate the propensity scores for matching and adjusting patient backgrounds included: sex, age, glycated hemoglobin (HbA1c), urine protein, eGFR, pre-index eGFR slope, and the use of concomitant medications such as biguanides, sulfonylureas, insulin, thiazolidinediones, α-glucosidase inhibitors, glinides, angiotensin-converting enzyme (ACE) inhibitors, angiotensin II receptor blockers, calcium channel blockers, diuretics, β-blockers, α-blockers, and statins.

We employed a DR learning framework to estimate the CATEs **[20]**, which represent the average treatment effect within specific subgroups defined by key baseline characteristics (e.g., sex, presence of proteinuria, or baseline eGFR). Understanding CATEs is crucial to identifying populations who may derive the most renoprotective benefit from a SGLT2 inhibitor, thus informing more personalized clinical decision-making. This approach mitigates the risk of model misspecification by combining the propensity score (PS) model with the outcome regression model, ensuring accurate estimates even if one of the models is misspecified **[21]**. All baseline variables were included as confounders in the PS, outcome regression, and final models to account for any potential imbalances and to identify subgroups with different CATEs. The dataset was randomly split into training (80%) and test (20%) sets. The training set was used for model development and hyperparameter tuning, while the test set evaluated model performance.

Models were developed in two stages. In the first stage, we estimated the predicted probability of receiving the medication of interest for each subject using a PS model with an inverse probability of treatment weighting (IPTW) approach. We also estimated the expected eGFR slope for both the SGLT2 inhibitor group and the DPP4 inhibitor group using an outcome regression model. Extreme gradient boosting (XGBoost) was used for the PS model and outcome regression model. Optimal hyperparameters were selected using cross-validated models with “GridSearchCV” or “RandomizedSearchCV” based on area under the curve (AUC) or root mean squared error (RMSE). In the second stage, we integrated the two models from the first stage into the final model to estimate treatment effects using “LinearDRLearner,” a supervised machine learning model developed in the EconML package **[22]**.

Treatment effects, encompassing both CATEs and predicted ITEs at the patient level, were reported as the mean differences in eGFR slopes between the two groups, accompanied by 95% CIs. A positive treatment effect value indicated a slower chronic eGFR decline in the SGLT2 inhibitor group compared with the DPP4 inhibitor group. To quantify the continuous spectrum of individual characteristic attributions and identify the variables associated with treatment heterogeneity, SHAPley Additive exPlanations (SHAP) analysis was applied to the trained framework. Furthermore, a decision tree was employed to identify the most important characteristics and to explore the variables associated with treatment effect heterogeneity, with at least 5% of the total study sample required in each leaf (subgroup). The decision tree model was trained to maximize the difference in treatment effects between subgroups, identifying patient subgroups with varying responses to the exposure regarding each renal outcome.

Statistical analyses were performed using SAS version 9.4 (SAS Institute Inc.), Python version 3.10.19 (Python Software Foundation).

## RESULTS

### Baseline Patient Characteristics

A total of 4588 patients were included in the analysis, comprising 1659 patients in the SGLT2 inhibitor group and 2929 patients in the DPP4 inhibitor group. At baseline, several clinical parameters were well-balanced between the cohorts (**Table 1**), most notably the pre-index eGFR slope (-1.7 mL/min/1.73 m²/year in both groups), HbA1c (7.3%), and the proportion of women (39-40%). However, baseline imbalances (defined as SMD > 0.1) were observed; compared to the DPP4 inhibitor group, patients initiating SGLT2 inhibitors were significantly younger (mean age 60.8 vs. 69.9 years) and had a slightly higher baseline eGFR (64.3 vs. 60.5 mL/min/1.73 m²). Furthermore, the SGLT2 inhibitor group exhibited a higher baseline utilization of cardiovascular and renoprotective medications, including ACE inhibitors (11.6% vs. 4.0%), angiotensin II receptor blockers (45.9% vs. 38.6%), and diuretics (30.4% vs. 17.9%), as well as β-blockers (28.1% vs. 14.2%) and statins (49.2% vs. 41.3%). The SGLT2 inhibitor group also had a higher prevalence of dipstick-positive proteinuria (32.9% vs. 29.2%).

**Table 1.** Baseline characteristics of patients with DKD in the SGLT2 inhibitor and DPP4 inhibitor groups.

| Variables |  | Overall<br>n=4588 | DPP4<br>inhibitor<br>n=2929 | SGLT2<br>inhibitor<br>n=1659 | SMD* | P value† |
| --- | --- | --- | --- | --- | --- | --- |
| Sex | Women | 1812 (39.5) | 1168 (39.9) | 644 (38.8) | 0.022 | 0.501 |
|  | Men | 2776 (60.5) | 1761 (60.1) | 1015 (61.2) |  |  |
| Age (years) |  | 66.6 (13.3) | 69.9 (11.5) | 60.8 (14.2) | 0.702 | <0.001 |
| Hemoglobin A1c (%) |  | 7.3 (1.4) | 7.3 (1.3) | 7.3 (1.6) | 0.057 | 0.07 |
| Urine Protein | Negative | 3188 (69.5) | 2075 (70.8) | 1113 (67.1) | 0.081 | 0.009 |
|  | Positive | 1400 (30.5) | 854 (29.2) | 546 (32.9) |  |  |
| eGFR (mL/min/1.73m<sup>2</sup>) |  | 61.9 (21.6) | 60.5 (21.4) | 64.3 (21.8) | 0.175 | <0.001 |
| eGFR slope (mL/min/1.73m<sup>2</sup>/year) |  | -1.7 (5.4) | -1.7 (5.9) | -1.7 (4.5) | 0.005 | 0.87 |
| **Medication** |  |  |  |  |  |  |
| Biguanides | No | 3611 (78.7) | 2351 (80.3) | 1260 (75.9) | 0.105 | <0.001 |
|  | Yes | 977 (21.3) | 578 (19.7) | 399 (24.1) |  |  |
| Sulfonylureas | No | 4213 (91.8) | 2663 (90.9) | 1550 (93.4) | 0.094 | 0.003 |
|  | Yes | 375 (8.2) | 266 (9.1) | 109 (6.6) |  |  |
| Insulin | No | 3524 (76.8) | 2253 (76.9) | 1271 (76.6) | 0.007 | 0.841 |
|  | Yes | 1064 (23.2) | 676 (23.1) | 388 (23.4) |  |  |
| Thiazolidinediones | No | 4353 (94.9) | 2784 (95.0) | 1569 (94.6) | 0.021 | 0.528 |
|  | Yes | 235 (5.1) | 145 (5.0) | 90 (5.4) |  |  |
| α-Glucosidase inhibitors | No | 4109 (89.6) | 2576 (87.9) | 1533 (92.4) | 0.15 | <0.001 |
|  | Yes | 479 (10.4) | 353 (12.1) | 126 (7.6) |  |  |
| Glinide | No | 4311 (94.0) | 2710 (92.5) | 1601 (96.5) | 0.175 | <0.001 |
|  | Yes | 277 (6.0) | 219 (7.5) | 58 (3.5) |  |  |
| angiotensin-converting<br>enzyme (ACE) inhibitors | No | 4278 (93.2) | 2811 (96.0) | 1467 (88.4) | 0.284 | <0.001 |
|  | Yes | 310 (6.8) | 118 (4.0) | 192 (11.6) |  |  |
| Angiotensin II receptor<br>blockers (ARB) | No | 2694 (58.7) | 1797 (61.4) | 897 (54.1) | 0.148 | <0.001 |
|  | Yes | 1894 (41.3) | 1132 (38.6) | 762 (45.9) |  |  |
| Calcium channel blockers | No | 2677 (58.3) | 1691 (57.7) | 986 (59.4) | 0.035 | 0.275 |
|  | Yes | 1911(41.7) | 1238 (42.3) | 673 (40.6) |  |  |
| Diuretics | No | 3559 (77.6) | 2405 (82.1) | 1154 (69.6) | 0.296 | <0.001 |
|  | Yes | 1029 (22.4) | 524 (17.9) | 505 (30.4) |  |  |
| **β-Blocker** | **No** | 3707 (80.8) | 2514 (85.8) | 1193 (71.9) | 0.346 | <0.001 |
|  | **Yes** | 881 (19.2) | 415 (14.2) | 466 (28.1) |  |  |
| **α-Blocker** | **No** | 4459 (97.2) | 2830 (96.6) | 1629 (98.2) | 0.099 | 0.003 |
|  | **Yes** | 129 (2.8) | 99 (3.4) | 30 (1.8) |  |  |
| **Statin** | **No** | 2562 (55.8) | 1719 (58.7) | 843 (50.8) | 0.159 | <0.001 |
|  | **Yes** | 2026 (44.2) | 1210 (41.3) | 816 (49.2) |  |  |

Values are presented as mean (SD) for continuous variables and n (%) for categorical variables.
\* SMD, standardized mean difference.
† P values were calculated using the Student's t-test for continuous variables and the chi-square test for categorical variables.
Abbreviations: DPP4, dipeptidyl peptidase-4; SGLT2, sodium-glucose cotransporter 2.

### Model Performance and Distribution of Treatment Effects

The XGBoost classifier, utilized for the propensity score model, demonstrated discriminative capability, achieving a mean validation AUC of 0.810 across five folds and a final test AUC of 0.817. Concurrently, the XGBoost regressor, employed for the outcome regression model for eGFR slope, exhibited stable error rates, yielding a mean validation RMSE of 18.65 mL/min/1.73m²/year and a final test RMSE of 17.72 mL/min/1.73m²/year.

We calculated the ITEs for the entire cohort. The distribution of these estimated treatment effects revealed an overall mean effect of 0.14 (95% CIs −0.86 to 1.15) mL/min/1.73m²/year on the chronic eGFR slope, suggesting a modest average renoprotective benefit favoring SGLT2 inhibitors over DPP4 inhibitors (**Figure S1**). When stratifying the estimated ITEs into deciles, a striking variation in treatment response emerged (**Figure S2**). To further investigate the clinical variables associated with treatment response, we evaluated the CATEs across predefined conventional clinical subgroups. When stratifying the population by baseline variables, formal interaction testing revealed no statistically significant heterogeneity across any subgroup (**Table S1**).

### Variables Associated with Treatment Heterogeneity via SHAP Analysis

To assess the variables associated with the ITEs, we utilized SHAP values derived from the Linear DR-learner model (**Figure 1**). A lower baseline eGFR, younger age, the presence of dipstick-positive proteinuria, insulin therapy, and female sex correlated with a positive SHAP value.

**Figure 1.**
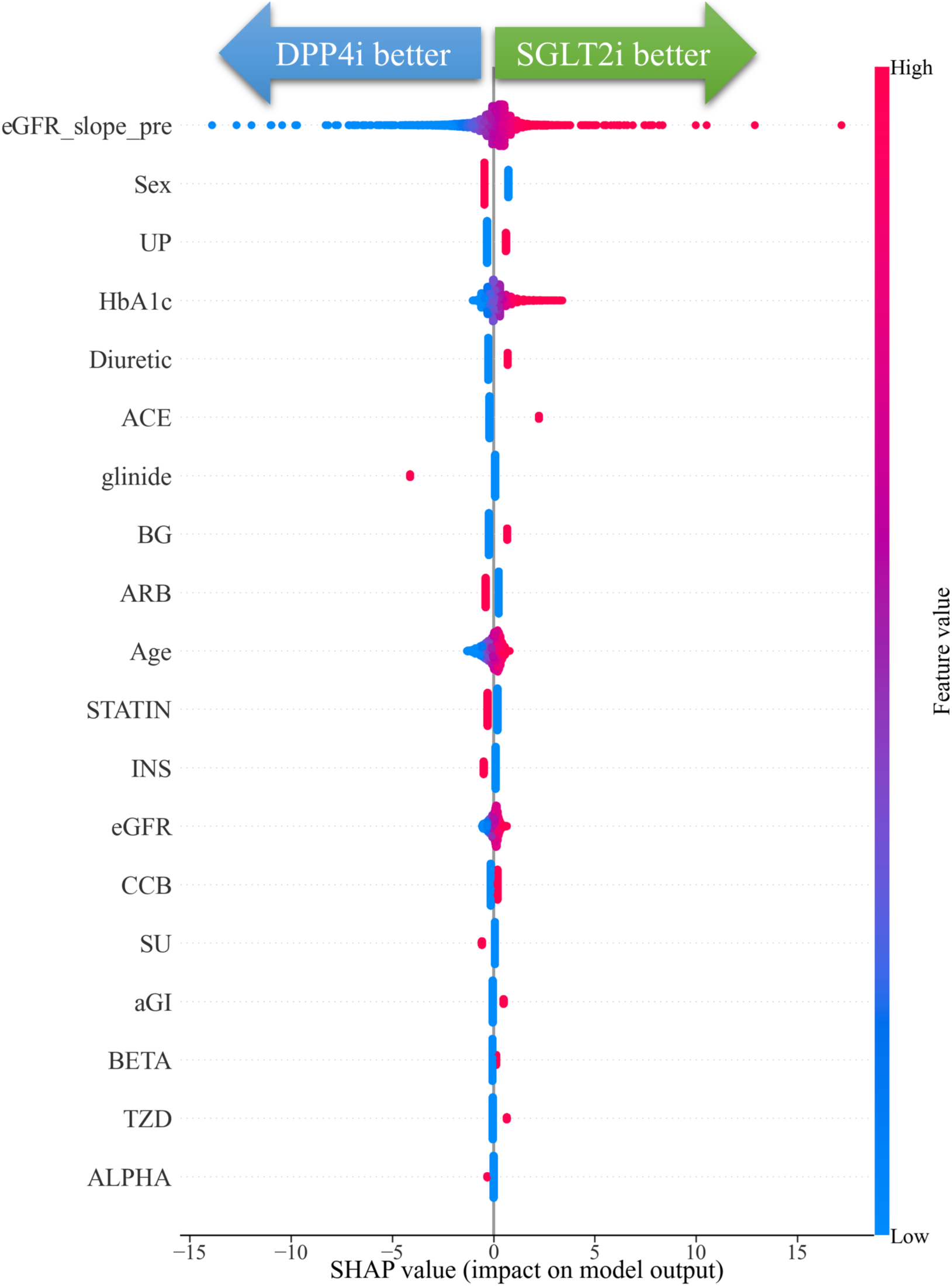
SHAP summary plot illuminating baseline feature attributions to individualized treatment effects on the chronic eGFR slope. The beeswarm plot ranks baseline clinical characteristics from top to bottom based on their magnitude of contribution to the treatment effect heterogeneity between SGLT2 inhibitors and DPP4 inhibitors. Each dot represents an individual patient. The x-axis denotes the SHAP value: positive values (<0) indicate an enhanced therapeutic response favoring SGLT2 inhibitors (SGLT2i better), whereas negative values (>0) represent a response favoring DPP4 inhibitors (DPP4i better). The color spectrum signifies the relative feature value, where red represents a high value and blue represents a low value. *Abbreviations:* ACE, angiotensin-converting enzyme inhibitor; aGI, *α*-glucosidase inhibitor; ALPHA, *α*-blocker; ARB, angiotensin II receptor blocker; BETA, *β*-blocker; BG, biguanide; CCB, calcium channel blocker; DPP4i, dipeptidyl peptidase-4 inhibitor; eGFR, estimated glomerular filtration rate; eGFR_slope_pre, pre-treatment eGFR trajectory; HbA1c, glycated hemoglobin; INS, insulin; SHAP, SHAPley Additive exPlanations; SGLT2i, sodium-glucose cotransporter 2 inhibitor; SU, sulfonylurea; TZD, thiazolidinedione; UP, urine protein.

### Multidimensional Patient Stratification via Treatment Effect Heterogeneity

Among patients not taking glinides, the ATE was positive at 0.39, indicating an overall preference for SGLT2 inhibitors (**Figure 2**). Within this group, those with a pre-index eGFR slope > −6.55 mL/min/1.73 m²/year who were concomitantly utilizing ACE inhibitors demonstrated a benefit from SGLT2 inhibitors, with an ATE of 2.95 (95% CI: −0.68 to 6.58) mL/min/1.73 m²/year. Conversely, patients with glinides who had a pronounced steep decline with a pre-index eGFR slope ≤ −4.48 mL/min/1.73 m²/year, and further sub-stratified to ≤ −14.36 mL/min/1.73 m²/year exhibited a negative ATE of −8.98 (95% CI: −16.11 to −1.85) mL/min/1.73m²/year.

**Figure 2.**
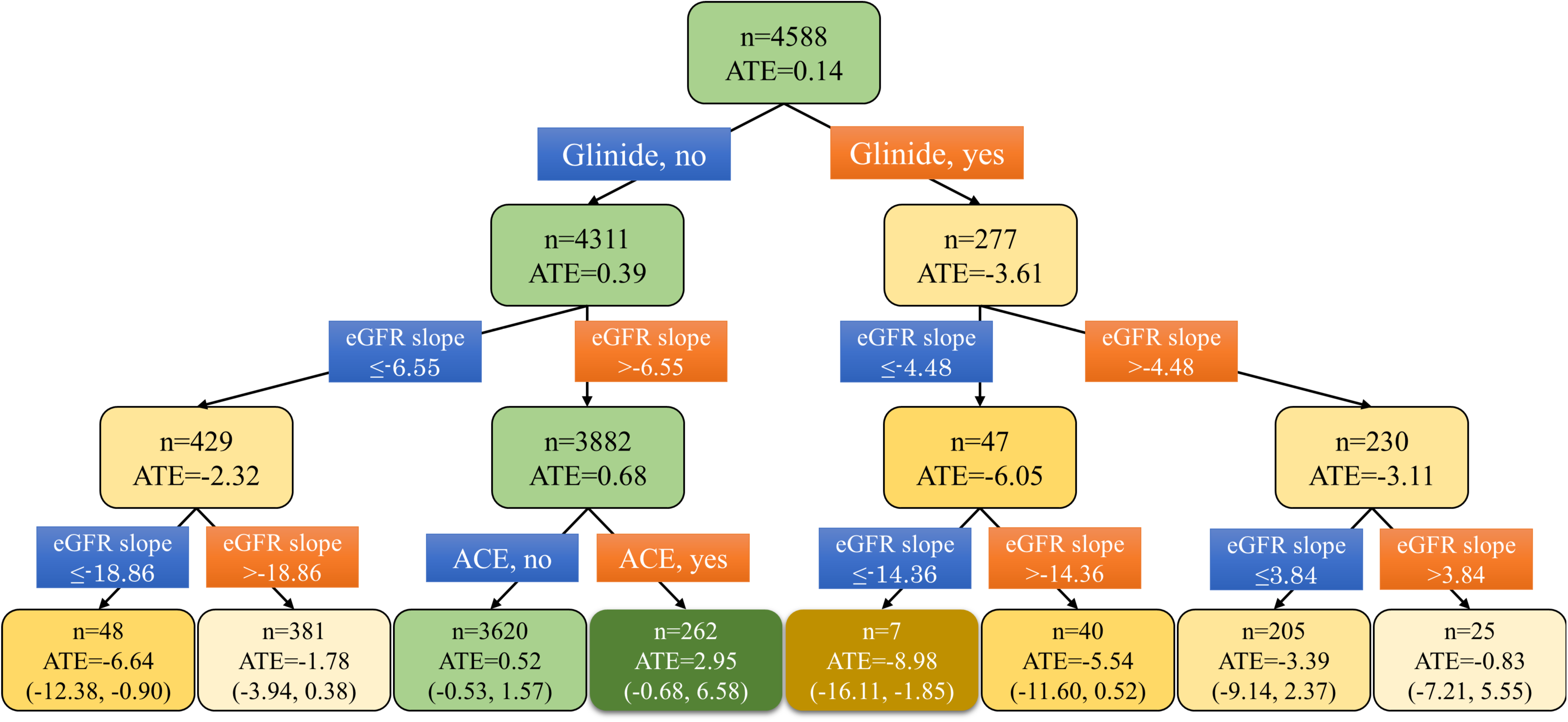
Causal decision tree partitioning heterogeneous treatment effects on the chronic eGFR slope. This tree diagram illustrates the sequential stratification of the study population (n=4558) based on variables associated with treatment effect heterogeneity. The model was trained to maximize treatment effect differentials between subgroups. The color gradient of the nodes reflects the differences in treatment effects: green nodes signify a positive treatment response favoring SGLT2 inhibitors, while orange/brown nodes signify a negative response favoring DPP4 inhibitors. Each node lists the sample size (*n*) and the estimated average treatment effect (ATE) on the chronic eGFR slope (mL/min/1.73 m²/year). Parentheses in the terminal leaves indicate the 95% confidence intervals. *Abbreviations:* ACE, angiotensin-converting enzyme inhibitor; ATE, average treatment effect; DPP4, dipeptidyl peptidase-4; eGFR, estimated glomerular filtration rate; SGLT2, sodium-glucose cotransporter 2.

### Secondary Outcomes (composite renal outcomes)

Using XGBoost classifiers, the PS model demonstrated discriminative capacity, achieving a mean training AUC of 0.945, a mean validation AUC of 0.806 across five folds, and a final test AUC of 0.817. Similarly, the outcome prediction model exhibited a mean training AUC of 0.894, a mean validation AUC of 0.834, and a final test AUC of 0.801. The distribution of the ITEs revealed an overall mean effect of −0.09 (95% CI: −0.11 to −0.08), suggesting an average 9% absolute risk reduction in the composite endpoint for patients receiving SGLT2 inhibitors compared to those receiving DPP4 inhibitors at the population level (**Figure S3**). However, visualizing the distribution of these treatment effects exposed profound underlying heterogeneity within the patient population (**Figure S4**). When stratifying the estimated ITEs into deciles, patients in the lowest decile (Decile 1) exhibited exceptionally strong negative treatment effect values, approaching an absolute risk reduction of nearly 50%, demonstrating a massive protective benefit from SGLT2 inhibitors. This risk reduction gradually attenuated across subsequent deciles but remained clinically meaningful and in favor of SGLT2 inhibitors through Decile 8. Conversely, patients in the highest decile (Decile 10) demonstrated positive treatment effects, suggesting that a distinct minority of the population might not derive structural renoprotection from SGLT2 inhibitors, or could potentially experience a marginally higher risk compared to DPP4 inhibitors.

SHAP values demonstrated that a lower baseline eGFR, younger age, the presence of dipstick-positive proteinuria, concomitant insulin therapy, and female sex correlated with negative SHAP values, indicating greater renoprotective efficacy of SGLT2 inhibitors compared with DPP4 inhibitors (**Figure 3**).

**Figure 3.**
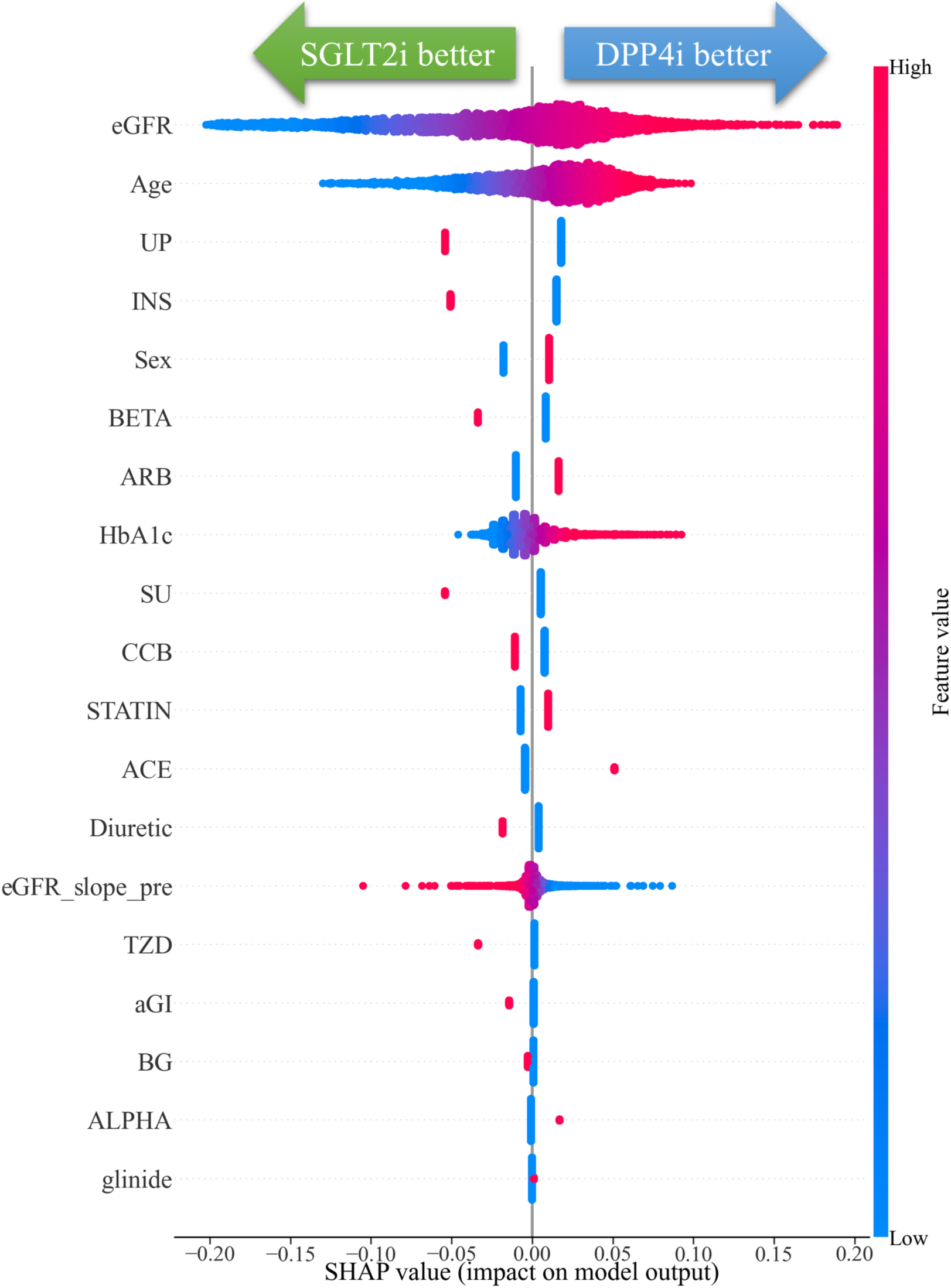
SHAP summary plot of baseline variables associated with treatment heterogeneity for the composite renal outcome comparing SGLT2 inhibitors versus DPP4 inhibitors. This plot illustrates the causal SHAP values for each baseline characteristic regarding the composite renal outcome, ordered vertically by overall model importance. Individual patients are depicted as single dots. The color gradient of the dots reflects the variations in baseline feature values (red for high, blue for low). The x-axis measures the directional influence on the individualized absolute risk reduction; negative SHAP values indicate a lower event risk with SGLT2 inhibitors (“SGLT2i better”), while positive values favor DPP4 inhibitors (“DPP4i better”). The wide, structured dispersion of dots unmasks the multidimensional nature of treatment effect heterogeneity across the population. ***Abbreviations:*** ACE, angiotensin-converting enzyme inhibitor; aGI, *α*-glucosidase inhibitor; ALPHA, *α*-blocker; ARB, angiotensin II receptor blocker; BETA, *β*-blocker; BG, biguanide; CCB, calcium channel blocker; DPP4i, dipeptidyl peptidase-4 inhibitor; eGFR, estimated glomerular filtration rate; eGFR_slope_pre, pre-treatment eGFR trajectory; HbA1c, glycated hemoglobin; INS, insulin; SHAP, SHAPley Additive exPlanations; SGLT2i, sodium-glucose cotransporter 2 inhibitor; SU, sulfonylurea; TZD, thiazolidinedione; UP, urine protein.

The root node algorithmically identified baseline eGFR as the primary bifurcating variable. Patients with an eGFR ≤ 42.4 mL/min/1.73m² had a greater ATE (−0.21), favoring SGLT2 inhibitors (**Figure 4**). Patients with a baseline eGFR 42.4 mL/min/1.73 m² and positive proteinuria, those who further exhibited advanced impairment ( 28.1 mL/min/1.73 m²) achieved a reduction for composite renal events with SGLT2 inhibitors (ATE = −0.28 [95% CI: −0.33 to −0.23]). Conversely, patients with eGFR > 42.4 mL/min/1.73m², further stratified to > 59.8 mL/min/1.73m², and negative proteinuria exhibited an ATE of −0.03 (95% CI: −0.05 to −0.01). Even within the non-proteinuric branch, the algorithm suggested a subgroup with eGFR <59.8 mL/min/1.73m² that still derived a renoprotective effect, yielding an ATE of −0.08 (95% CI: −0.10 to −0.06).

**Figure 4.**
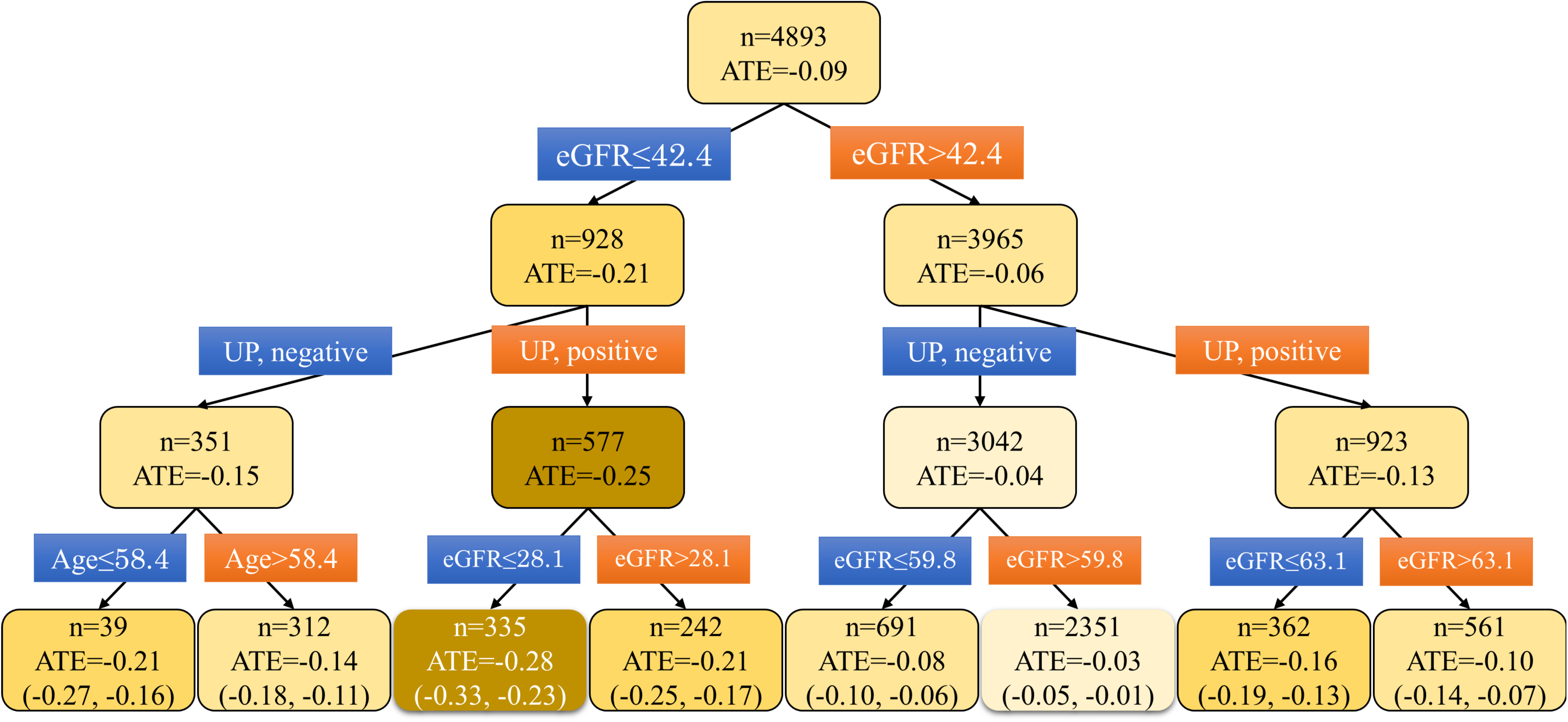
Causal decision tree partitioning heterogeneous absolute risk reductions for the composite renal outcome comparing SGLT2 inhibitors versus DPP4 inhibitors. This tree illustrates the sequential stratification of the patient population based on baseline characteristics associated with treatment effect heterogeneity in the composite renal outcome. The model was trained to maximize differences in treatment effects between subgroups. The color gradient of the nodes reflects the differences in treatment effects: because negative values represent absolute risk reduction for a clinical event, darker orange/brown shades signify a more pronounced renoprotective response favoring SGLT2 inhibitors over DPP4 inhibitors. Each node lists the sample size (*n*) and the estimated average treatment effect. Parentheses in the terminal leaves indicate the 95% confidence intervals. ***Abbreviations:*** ATE, average treatment effect; DPP4, dipeptidyl peptidase-4; eGFR, estimated glomerular filtration rate; SGLT2, sodium-glucose cotransporter 2; UP, urine protein.

## DISCUSSION

Using a doubly robust causal ML framework in the J-CKD-DB-Ex registry, we demonstrated heterogeneous treatment effects for SGLT2 inhibitors compared to DPP4 inhibitors across both functional and structural renal endpoints. Our causal SHAP analysis quantified individual characteristic attributions as a continuous spectrum, revealing that dipstick-positive proteinuria, elevated baseline HbA1c, and a less rapid pre-treatment eGFR decline were consistently associated with a therapeutic preference for SGLT2 inhibitors over DPP4 inhibitors across both endpoints.

For the chronic eGFR slope, the causal decision tree identified non-glinide users as a critical bifurcating pathway favoring SGLT2 inhibitors over DPP-4 inhibitors. Non-glinide users may represent an insulin-resistant, volume-expanded phenotype characterized by a well-preserved renal functional reserve. At the cellular level, these proximal tubular cells possess sufficient metabolic resilience to tolerate the expected acute hemodynamic dip and transient osmotic diuresis expected upon SGLT2 inhibitor initiation by activating adaptive pathways, such as autophagy and mitochondrial metabolic shifts.**[23]** Consequently, they might have the long-term chronic benefits of SGLT2 inhibition.

Within the non-glinide group, patients with a pre-index eGFR slope 6.55 mL/min/1.73 m²/year were further bifurcated by concomitant ACE inhibitor use, isolating a phenotype that derived significantly greater chronic eGFR slope preservation from SGLT2 inhibitors over DPP4 inhibitors. This reflects an intrarenal microvascular synergy where RAAS inhibitor-induced efferent vasodilation intersects with SGLT2 inhibitor-mediated afferent vasoconstriction, ultimately optimizing intraglomerular pressure and reducing tubulointerstitial barotrauma. **[24, 25]**

Conversely, caution is warranted when initiating SGLT2 inhibitors in patients with a rapidly declining pre-treatment eGFR trajectory. Among glinide users with a pre-treatment eGFR slope −4.48 mL/min/1.73 m²/year, those whose decline further culminated in a pre-index reduction of −14.36 mL/min/1.73 m²/year demonstrated a treatment effect that inverted to favor DPP4 inhibitors. This pattern was similar to non-glinide users with extreme pre-treatment eGFR slope drops. In these potentially hypoxic, ATP-depleted kidneys, the acute fluid shifts and volume shock induced by SGLT2 inhibitors might trigger ischemic re-injury. In contrast, hemodynamically neutral DPP4 inhibitors provide a safe clinical alternative by maintaining systemic and intrarenal microvascular stability.

For the composite renal endpoint, the causal tree prioritized structural severity, selecting baseline eGFR≤ 42.4 mL/min/1.73m²/year as the single root bifurcating variable. Within this arm, patients with eGFR ≤ 28.1 mL/min/1.73m²/year and concomitant proteinuria had a 28% absolute risk reduction with SGLT2 inhibitors over DPP4 inhibitors. This aligns with post-hoc evidence from the DAPA-CKD and EMPA-KIDNEY trials, suggesting that SGLT2 inhibitors significantly slow chronic decline and prevent hard endpoints even in Stage 4 CKD, especially with baseline proteinuria. **[26, 27]**

Finally, among patients without proteinuria, the algorithm isolated a large subgroup with eGFR≤ 59.8 mL/min/1.73m²/year that achieved a 8% absolute risk reduction with SGLT2 inhibitors. Because non-proteinuric DKD lacks the diagnostic warning of overt proteinuria, these early decliners are frequently under-treated. Our finding justifies early, proactive SGLT2 inhibitor initiation, regardless of proteinuria status, to optimize long-term nephroprotection. **[28]**

Several limitations of this study must be acknowledged. First, its retrospective registry design introduces potential unmeasured confounding factors, including blood pressure, patient compliance, and dietary habits. Second, medication exposure was assessed at baseline, without modeling subsequent discontinuations or cross-overs as time-varying exposures. Third, the findings from this Japanese cohort may lack generalizability to Western populations with distinct metabolic and macrovascular phenotypes. Finally, isolating specific multi-dimensional pathways reduced the sample size within the extreme tree nodes, meaning the point estimates for these subgroups should be validated in larger studies.

In conclusion, we demonstrated the utility of causal ML in advancing precision medicine by mapping heterogeneous treatment effects across functional and structural renal endpoints in DKD patients. Crucially, while the population-level average effect showed a modest 9% risk reduction in the composite renal outcome favoring SGLT2 inhibitors over DPP4 inhibitors, our framework successfully unmasked a hidden 28% absolute risk reduction subgroup for this same endpoint, demonstrating a methodological advantage over traditional 1:1 propensity score matching. Ultimately, these data-driven insights prove that rather than adhering to a uniform prescribing paradigm, therapeutic selection should be precisely tailored to a patient’s specific clinical profile and underlying pathophysiology. Evaluating characteristics, including proteinuria status, concomitant ACE inhibitor use, and pre-treatment eGFR trajectories allows clinicians to strategically leverage the distinct renoprotective mechanisms of SGLT2 and DPP4 inhibitors to achieve optimized patient outcomes.

## Disclosure

The authors declare no conflicts of interest.

## Data Availability

All data produced in the present study are available upon reasonable request to the authors.

## Acknowledgements

This work was supported by AMED under Grant Number JP25ek0109812.

