## Supplemental Table and Figures for "Individual-Level Counterfactual Analysis of SGLT2 Inhibitors Versus DPP4 Inhibitors in Diabetic Kidney Disease Using Causal Machine Learning"

### SUPPLEMENTARY TABLE

**Table S1. Conditional average treatment effects on the chronic eGFR slope comparing SGLT2 inhibitors versus DPP4 inhibitors, stratified by baseline characteristics**

| Variables | | n | ATE* | 95%CI | P value |
|---|---|---|---|---|---|
| **Sex** | **Female** | 1812 | 0.71 | (-0.84, 2.27) | 0.37 |
| | **Male** | 2776 | -0.23 | (-1.54, 1.08) | 0.735 |
| | **Difference** | | 0.94 | (-1.10, 2.97) | 0.366 |
| **Age** | **<70 years** | 2424 | 0.03 | (-1.21, 1.26) | 0.968 |
| | **≥70 year** | 2164 | 0.28 | (-1.08, 1.63) | 0.69 |
| | **Difference** | | -0.25 | (-2.08, 1.58) | 0.789 |
| **Hemoglobin A1c** | **<7.0%** | 1992 | -0.28 | (-1.61, 1.05) | 0.678 |
| | **≥7.0%** | 2596 | 0.47 | (-0.83, 1.76) | 0.478 |
| | **Difference** | | -0.75 | (-2.60, 1.10) | 0.428 |
| **Urine Protein** | **Negative** | 3188 | 0.13 | (-1.10, 1.36) | 0.838 |
| | **Positive** | 1400 | 0.18 | (-1.54, 1.89) | 0.839 |
| | **Difference** | | -0.05 | (-2.16, 2.06) | 0.964 |
| **eGFR** | **<60 mL/min/1.73m$^2$** | 1789 | -0.43 | (-1.85, 0.99) | 0.556 |
| | **≥60 mL/min/1.73m$^2$** | 2799 | 0.51 | (-0.65, 1.67) | 0.391 |
| | **Difference** | | -0.94 | (-2.77, 0.90) | 0.318 |
| **eGFR slope** | **<-1.0 mL/min/1.73m$^2$/year** | 2420 | -0.59 | (-1.79, 0.61) | 0.333 |
| | **≥-1.0 mL/min/1.73m$^2$/year** | 2168 | 0.96 | (-0.21, 2.14) | 0.108 |
| | **Difference** | | -1.55 | (-3.23, 0.12) | 0.069 |
| **Medication** | | | | | |
| **Biguanides** | **No** | 3611 | -0.06 | (-1.22, 1.09) | 0.916 |
| | **Yes** | 977 | 0.9 | (-1.08, 2.89) | 0.372 |
| | **Difference** | | -0.97 | (-3.26, 1.33) | 0.41 |
| **Sulfonylureas** | **No** | 4213 | 0.13 | (-0.94, 1.19) | 0.814 |
| | **Yes** | 375 | 0.32 | (-2.49, 3.13) | 0.822 |
| | **Difference** | | -0.19 | (-3.20, 2.81) | 0.899 |
| **Insulin** | **No** | 3524 | 0.19 | (-0.90, 1.27) | 0.737 |
| | **Yes** | 1064 | 0 | (-2.40, 2.40) | 0.999 |
| | **Difference** | | 0.19 | (-2.45, 2.82) | 0.89 |
| **Thiazolidinediones** | **No** | 4353 | 0.11 | (-0.92, 1.15) | 0.829 |
| | **Yes** | 235 | 0.69 | (-3.29, 4.67) | 0.735 |

| | | | | | |
|---|---|---|---|---|---|
| | **Difference** | | -0.57 | (-4.69, 3.54) | 0.785 |
| **α-Glucosidase inhibitors** | **No** | 4109 | 0.15 | (-0.93, 1.23) | 0.784 |
| | **Yes** | 479 | 0.08 | (-2.57, 2.73) | 0.952 |
| | **Difference** | | 0.07 | (-2.79, 2.93) | 0.962 |
| **Glinide** | **No** | 4311 | 0.38 | (-0.62, 1.39) | 0.452 |
| | **Yes** | 277 | -3.61 | (-9.38, 2.16) | 0.22 |
| | **Difference** | | 3.99 | (-1.86, 9.85) | 0.181 |
| **angiotensin-converting enzyme (ACE) inhibitors** | **No** | 4278 | -0.03 | (-1.07, 1.02) | 0.962 |
| | **Yes** | 310 | 2.47 | (-1.16, 6.10) | 0.182 |
| | **Difference** | | -2.5 | (-6.27, 1.28) | 0.195 |
| **Angiotensin II receptor blockers (ARB)** | **No** | 2694 | 0.39 | (-0.99, 1.77) | 0.575 |
| | **Yes** | 1894 | -0.21 | (-1.65, 1.22) | 0.771 |
| | **Difference** | | 0.61 | (-1.38, 2.60) | 0.55 |
| **Calcium channel blockers** | **No** | 2677 | 0.13 | (-1.21, 1.47) | 0.848 |
| | **Yes** | 1911 | 0.16 | (-1.36, 1.68) | 0.835 |
| | **Difference** | | -0.03 | (-2.05, 1.99) | 0.976 |
| **Diuretics** | **No** | 3559 | 0 | (-1.12, 1.11) | 0.994 |
| | **Yes** | 1029 | 0.66 | (-1.64, 2.95) | 0.575 |
| | **Difference** | | -0.66 | (-3.21, 1.89) | 0.612 |
| **β-Blocker** | **No** | 3707 | 0.04 | (-1.07, 1.15) | 0.944 |
| | **Yes** | 881 | 0.58 | (-1.80, 2.96) | 0.632 |
| | **Difference** | | -0.54 | (-3.16, 2.08) | 0.686 |
| **α-Blocker** | **No** | 4459 | 0.16 | (-0.85, 1.18) | 0.751 |
| | **Yes** | 129 | -0.6 | (-6.27, 5.08) | 0.837 |
| | **Difference** | | 0.76 | (-5.00, 6.52) | 0.796 |
| **Statin** | **No** | 2562 | 0.26 | (-1.20, 1.72) | 0.727 |
| | **Yes** | 2026 | 0 | (-1.33, 1.33) | 0.996 |
| | **Difference** | | 0.26 | (-1.71, 2.24) | 0.794 |

Treatment effects were estimated using the doubly robust "LinearDRLearner" framework. A positive ATE value signifies a slower chronic eGFR decline in the SGLT2 inhibitor group compared with the DPP4 inhibitor group. The rows labeled "Difference" represent the estimated differential treatment effects between the corresponding stratified subgroups (e.g., Female vs. Male). The accompanying P values function as formal statistical tests for interaction, evaluating whether the therapeutic effects between the two drug classes significantly varies across the strata. No conventional clinical subgroup met the threshold for statistically significant heterogeneity ($P < 0.05$).

Abbreviations: ACE, angiotensin-converting enzyme; ATE, average treatment effect (or conditional average treatment effect within specific strata); ARB, angiotensin II receptor blocker; CI, confidence interval; eGFR, estimated glomerular filtration rate; HbA1c, glycated hemoglobin; SGLT2, sodium-glucose cotransporter 2; DPP4, dipeptidyl peptidase-4.* ATE, average treatment effect.

### SUPPLEMENTARY FIGURES

**Figure S1. Distribution of estimated individualized treatment effects on the chronic eGFR slope across the study population**

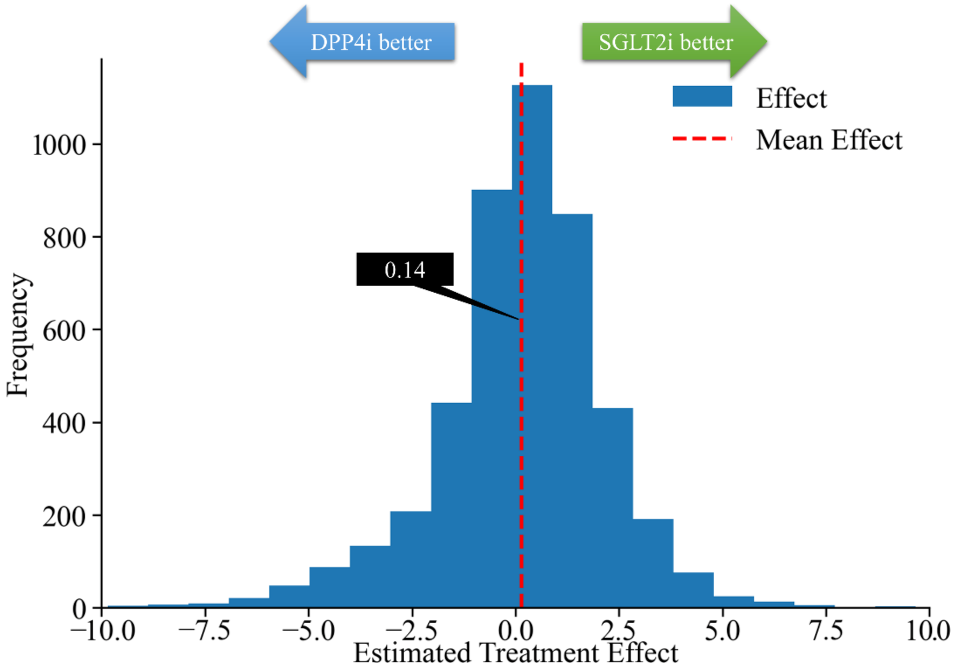


This histogram depicts the variations in estimated individualized treatment effects on the chronic eGFR slope, comparing SGLT2 inhibitors versus DPP4 inhibitors. The vertical red dashed line indicates the population-level mean effect of 0.14 mL/min/1.73 $m^2$/year. Positive values on the x-axis correspond to an enhanced therapeutic response with SGLT2 inhibitors ("SGLT2i better"), while negative values signify a more favorable trajectory with DPP4 inhibitors ("DPP4i better"). The wide spread of the distribution unmasks substantial underlying treatment heterogeneity across the patient population.

Abbreviations: DPP4i, dipeptidyl peptidase-4 inhibitor; eGFR, estimated glomerular filtration rate; SGLT2i, sodium-glucose cotransporter 2 inhibitor.Figure S2. Distribution of treatment effect

**Figure S2. Decile stratification of individualized treatment effects on the chronic eGFR slope comparing SGLT2 inhibitors versus DPP4 inhibitors**

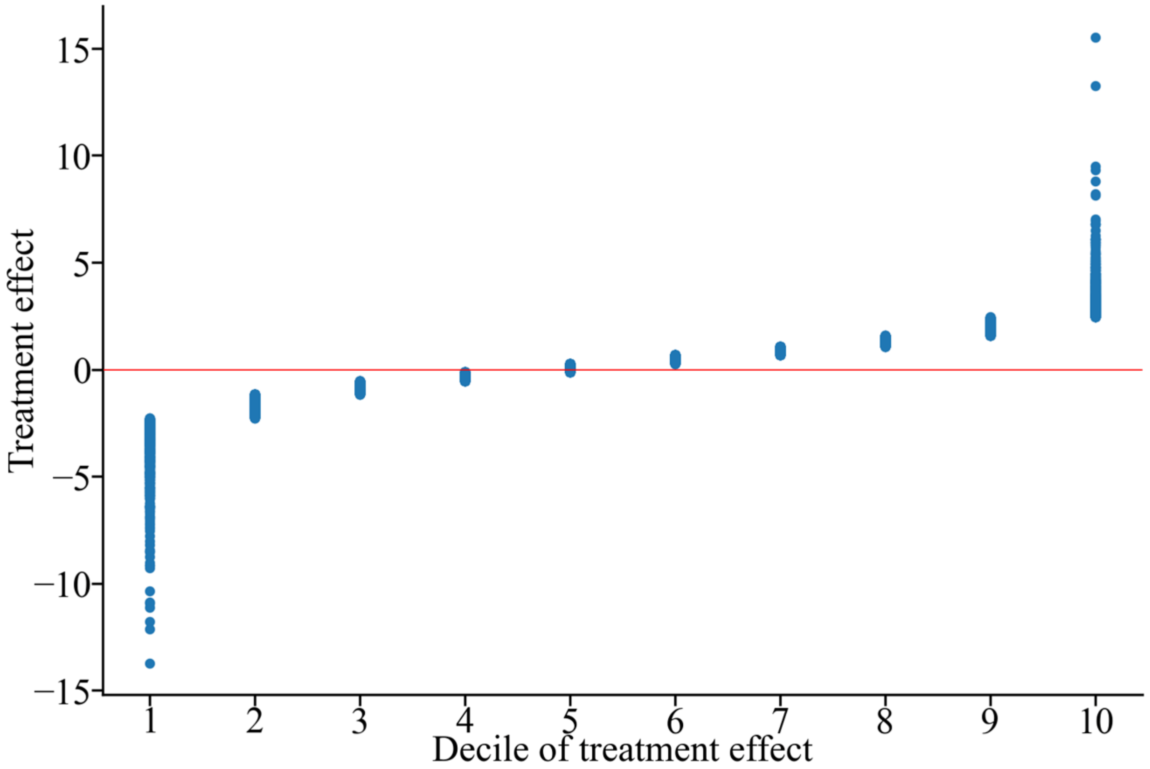


This plot presents the cohort divided into deciles based on the magnitude and direction of the predicted individualized treatment effects on the chronic eGFR slope, comparing SGLT2 inhibitors versus DPP4 inhibitors. The horizontal red line indicates a treatment effect of zero. Points above the line represent individual patient responses favoring SGLT2 inhibitors, while points below the line favor DPP4 inhibitors. The progressive upward trend from Decile 1 to Decile 10 demonstrates a wide, continuous spectrum of clinical responsiveness between the two medication classes.

*Abbreviations:* DPP4i, dipeptidyl peptidase-4 inhibitor; eGFR, estimated glomerular filtration rate; SGLT2i, sodium-glucose cotransporter 2 inhibitor.

**Figure S3. Frequency distribution of individualized absolute risk reduction for the composite renal outcome comparing SGLT2 inhibitors versus DPP4 inhibitors**

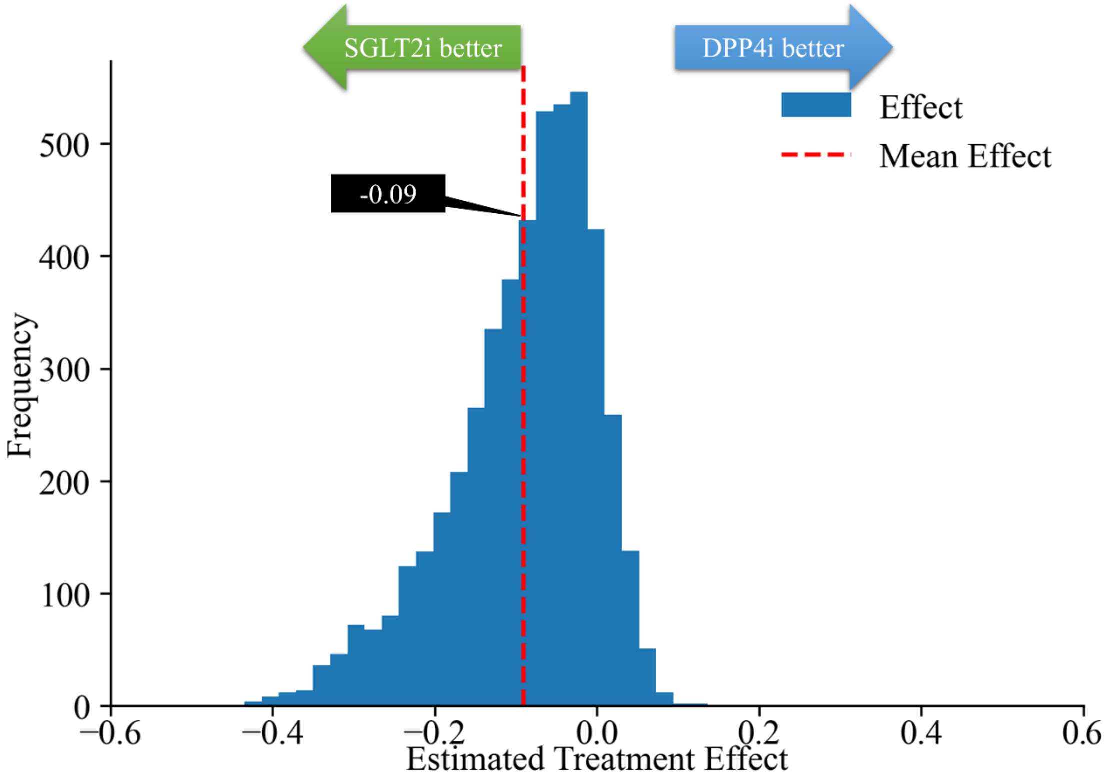


This histogram depicts the variation in predicted individualized treatment effects for the composite renal outcome (sustained ≥50% eGFR decline or end-stage kidney disease), comparing SGLT2 inhibitors versus DPP4 inhibitors. The x-axis reflects the absolute risk reduction, where negative values indicate a lower event risk with SGLT2 inhibitors ("SGLT2i better", green arrow) and positive values favor DPP4 inhibitors ("DPP4i better", blue arrow). The vertical red dashed line marks the population average treatment effect of -0.09 (9% absolute risk reduction). The wide, left-skewed spread of individual values unmasks deep heterogeneity in therapeutic response across the patient cohort.

*Abbreviations:* DPP4i, dipeptidyl peptidase-4 inhibitor; eGFR, estimated glomerular filtration rate; SGLT2i, sodium-glucose cotransporter 2 inhibitor.

**Figure S4. Decile stratification of individualized absolute risk reduction for the composite renal outcome comparing SGLT2 inhibitors versus DPP4 inhibitors**

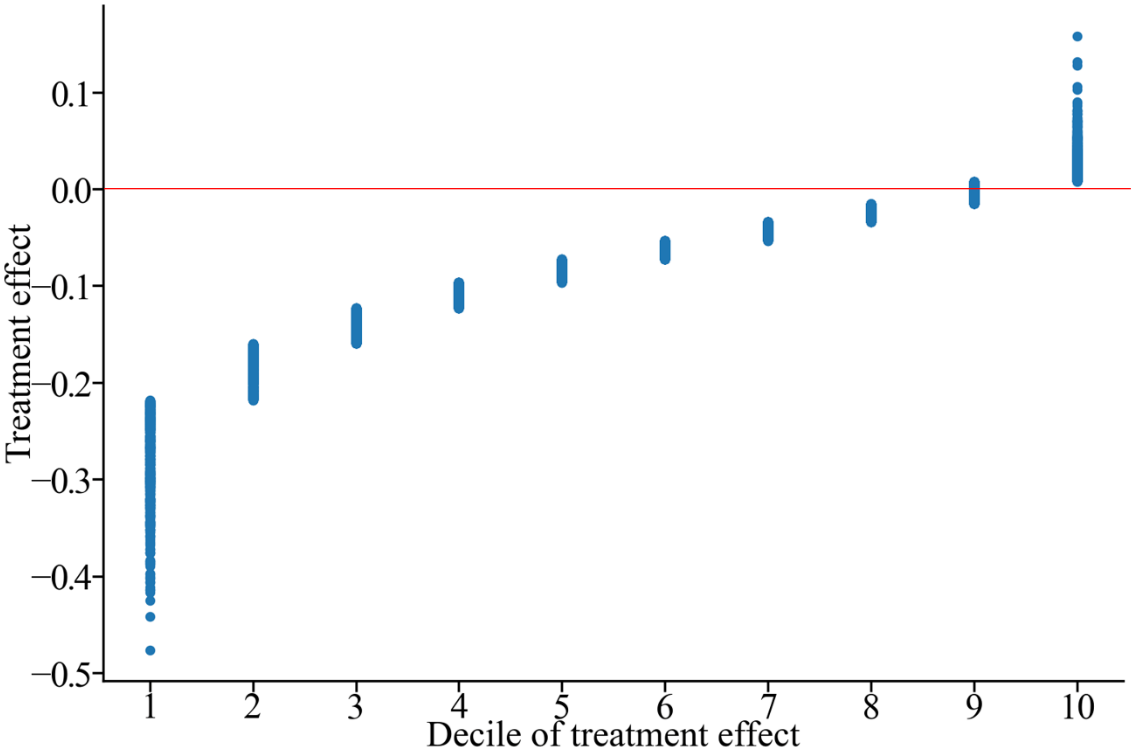


This plot presents the study population divided into deciles based on the magnitude and direction of the predicted individualized absolute risk reduction for the composite renal outcome (sustained ≥50% eGFR decline or end-stage kidney disease), comparing SGLT2 inhibitors versus DPP4 inhibitors. Negative values on the y-axis indicate a lower risk of experiencing the composite event with SGLT2 inhibitors, highlighted by Decile 1 where the risk reduction approaches 50%. Positive values in Decile 10 signify a subset of patients favoring DPP4 inhibitors. The progressive upward trend across the deciles highlights the deep heterogeneity in clinical responsiveness between the two drug classes.

*Abbreviations:* DPP4i, dipeptidyl peptidase-4 inhibitor; eGFR, estimated glomerular filtration rate; SGLT2i, sodium-glucose cotransporter 2 inhibitor.
